# Comparative Effectiveness Of Bedaquiline On One-Year Mortality In Rifampicin-Resistant Tuberculosis: A Target Trial Emulation

**DOI:** 10.1101/2024.08.23.24312479

**Authors:** Miriam Ngarega, Felex Ndebele, Pulane Segwaba, Sthabiso Bohlela, Zandile Sibeko, Leole Setlhare, Lesly E Scott, Wendy Stevens, Boitumelo Fanampe, Salome Charalambous, Gavin Churchyard, Annelies Van Rie

## Abstract

**Background:** Three phase II clinical trials generated the evidence for recommending bedaquiline for the treatment of rifampicin-resistant tuberculosis (RR-TB). These trials were not powered to assess the effect of bedaquiline on mortality. Observational studies reported lower mortality in patients treated with bedaquiline-containing regimens but did not fully account for differences between patients who did and did not receive bedaquiline in the real world.

**Methods:** Using data from two studies on 622 patients, of whom 195 initiated a bedaquiline-containing regimen, we applied rigorous causal inference by emulating a trial that would randomize patients diagnosed with RR-TB by the Xpert MTB/RIF assay to a bedaquiline-containing regimen or a non-bedaquiline-containing regimen. We used multiple imputation to address missing data, inverse probability of treatment weighting (IPTW) to emulate randomized assignment and estimated the odds of one-year mortality using a marginal structural logistic model.

**Results:** By using IPTW, we achieved conditional exchangeability for observed differences in age, gender, HIV status, *Mycobacterium tuberculosis* resistance pattern, and history of tuberculosis treatment between patients who did or did not initiate a bedaquiline-containing regimen. By emulating the design of a randomized trial, we found that had all patients been treated with a bedaquiline-containing regimen, there would have been a 67% reduction in the odds of one-year mortality compared to when none of the patients initiated a bedaquiline-containing regimen (OR: 0·33, 95%CI: 0·19-0·59)

**Conclusion:** By emulating a randomized trial using real-world data, our results demonstrate that the initiation of a bedaquiline-containing regimen causes a 67% reduction in the odds of one-year mortality.

**Key message:** We assessed the causal effect of initiating a bedaquiline-containing regimen compared to a non-bedaquiline-containing regimen on one-year mortality. We found that a bedaquiline-containing regimen causes a 67% reduction in the odds of one-year mortality, underscoring the need for expanded access to such effective regimens.

## 1. Introduction

The World Health Organisation (WHO) reported that 410 000 people developed rifampicin-resistant tuberculosis (RR-TB) in 2022, of which less than half (175 650) started RR-TB treatment. Despite important progress made in the diagnosis and treatment of RR-TB, the global treatment success rate among those who start treatment was only 63% and an estimated 160 000 died from the disease (1). RR-TB thus remains a critical public health challenge that requires global attention and action.

Approval of bedaquiline in 2012 by the Food and Drug Administration (FDA) paved the way for shorter, all-oral regimen for RR-TB (2). However, the two pivotal trials (3, 4) that led to the approval of bedaquiline observed a higher mortality rate in the patients randomized to bedaquiline. A meta-analysis combining the results of the two phase II trials found that patients in the bedaquiline arms had a four times higher risk of mortality compared to the placebo group (5). After the FDA approval, the WHO issued a conditional recommendation on the use of bedaquiline where it would be offered through compassionate access programmes under operational research conditions. In December 2012, South Africa was the first country to implement the Bedaquiline Clinical Access Programme (BCAP). The outcome of the Grading Recommendations, Assessment, Development and Evaluation (GRADE) process on the global use of bedaquiline in 2016 upheld the prevailing conditional WHO recommendations (6). In 2019, the WHO listed bedaquiline as a group A drug and made a strong recommendation that bedaquiline should be included in longer (18-month) RR-TB regimens for adults and a conditional recommendation that a shorter (9-12 months) bedaquiline-containing regimen may be used instead of longer regimens in selected patients (7). In a 2022 update, the WHO conditionally recommended the use of a 6-month treatment regimen composed of bedaquiline, pretomanid, linezolid and moxifloxacin (BPaLM) for RR-TB rather than the 9-month or 18-month regimen (8). Since 2019, several countries have increasingly adopted the use of bedaquiline because of its proven efficacy.

An observational study assessing the effect of bedaquiline on mortality reported a reduction in the hazard of mortality when used for treatment of patients with RR-TB, multi-drug resistant TB (MDR-TB) and extensively drug-resistant tuberculosis (XDR-TB) (9). The study population in the study was strictly those under the BCAP, thus not generalizable. Another study done after the national rollout of bedaquiline in South Africa also observed a reduced risk of mortality (10). While this study aimed to overcome substantial differences between patients treated with bedaquiline and those treated with injectables, the use of propensity score matching results in the exclusion of eligible participants who are not exactly matched as is sensitive to residual confounding depending on the caliper distance chosen. The trial Standard Treatment Regimen of Anti-tuberculosis Drugs for Patients With MDR-TB II (STREAM 2), which was the first phase III to evaluate the efficacy and safety of bedaquiline observed a higher mortality rate among the patients randomized to the 9-month oral regimen containing bedaquiline compared to those randomized to the 9-month control regimen without bedaquiline (11). A meta-analysis of randomised and non-randomized studies found inconsistent results with randomized studies showing an increased risk of mortality whereas non-randomized studies showed a reduced risk of mortality (12). The question of whether bedaquiline increases or reduces is therefore still uncertain. Answering this question would require a well-designed, randomized phase III clinical trial powered to detect a significant difference in mortality, but this is no longer ethically possible.

The target trial emulation approach has been proposed as a methodology to answer causal questions using observational data when a clinical trial cannot be performed (13). The approach allows for clear articulation of causal questions which alleviates (some of) the biases typically overlooked in standard analyses of observational studies. In line with the recent guidance on how causal inferences can be made from observational data (14), we applied the target trial emulation approach to assess the causal effect of initiating a bedaquiline-containing regimen compared to a non-bedaquiline-containing regimen on one-year mortality in patients diagnosed with RR-TB.

## 2. Methods

### 2.1. Specification of the target trial

The hypothetical target trial would enrol adult patients (≥ 18 years) diagnosed with RR-TB by the Xpert MTB/RIF (Xpert) assay and initiated on RR-TB treatment. Eligible patients would be randomly assigned to a bedaquiline-containing RR-TB regimen or a non-bedaquiline-containing RR-TB regimen on the day of treatment initiation. The outcome of interest would be death in the first year after randomization. Each patient would be followed up from the time of randomization until death or one year of follow-up, whichever occurs first. The causal contrast of interest is the intention-to-treat effect, that is, comparing the probability of mortality had all patients initiated a bedaquiline-containing regimen with the probability of mortality had none of the patients initiated a bedaquiline-containing regimen, as they were randomized.

### 2.2. Data sources and emulation of the target trial

#### 2.2.1. Description of the data sources

We used data from two studies: ‘Evaluating the impact of Xpert on Tuberculosis-RIFampicin Resistance’ (EXIT-RIF) and ‘Sequencing Mycobacteria and Algorithm-determined resistant tuberculosis treatment trial’ (SMARTT) trial to emulate the target trial. EXIT-RIF was a prospective observational cohort study that evaluated the phased implementation of the Xpert assay in South Africa. This study recruited patients from January 2012 to December 2013. All patients were diagnosed and treated in the public sector by routine staff (15). None of the patients received a bedaquiline-containing regimen. The EXIT-RIF study population diagnosed by the Xpert assay can be viewed as the control arm of the target trial. The phase IV pragmatic SMARTT trial recruited patients from September 2021 to February 2023 to evaluate whole-genome sequencing (WGS)-guided treatment recommendation. Being pragmatic, the patients were also treated under programmatic conditions in the public sector by routine staff. The trial recruited patients from September 2021 to February 2023 (16). All patients received a bedaquiline-containing regimen and can therefore be viewed as the treatment arm of the target trial. Both studies collected extensive and accurate data on the treatments patients received, covariates and the outcome of one-year all-cause mortality.

#### 2.2.2. Emulation of the target trial

The eligibility criteria for the emulated trial were the same as that of the target trial (**Table 1**). In the emulation, patients were assigned to the treatment strategy that was consistent with their data at baseline. Patients in the SMARTT study were started on a short (9-month) regimen consisting of bedaquiline, clofazimine, delamanid, levofloxacin, pyrazinamide and terizidone or a long (18-24 months) regimen bedaquiline-containing regimen. Regimens were individualised based on the results of standard-of-care drug susceptibility tests or WGS results. Even after individualization, all patients continued to receive a bedaquiline-containing regimen. According to the guidelines at the time, patients in the EXIT-RIF study started on a short (9-month) regimen consisting of amikacin, moxifloxacin, ethionamide, terizidone, pyrazinamide and isoniazid or a long regimen (18-24 month) regimen. Regimens were individualized based on DST results. None of these patients received bedaquiline even after individualization.

**Table 1:** Specification of the target trial of comparative effectiveness of bedaquiline on one-year mortality and how it will be emulated using data from EXIT-RIF and SMARTT.

| Protocol element | Target trial protocol | Emulation using data from EXIT-RIF and SMARTT studies |
| --- | --- | --- |
| **Eligibility criteria** | Adult patients ($\geq 18$ years) with rifampicin-resistant tuberculosis (RR-TB) diagnosed by the Xpert-MTB/RIF assay and initiated on treatment for drug-resistant tuberculosis will be eligible for enrolment in the trial. | Same as the target trial. |
| **Treatment strategies** | 1. Initiate RR-TB treatment with a bedaquiline-containing regimen.<br>2. Initiate RR-TB treatment with a non-bedaquiline-containing regimen (either amikacin or capreomycin). | Same as the target trial. |
| **Treatment assignment** | The patients will be randomly assigned to either treatment strategy at the start of treatment and will be aware of the strategy they have been assigned to. | Patients will be assigned at baseline to the treatment strategy with which their data are consistent with. To emulate randomization, inverse probability of treatment weighting was used to adjust for the following baseline confounders identified by a DAG: age, sex, weight, HIV status, history of TB treatment, haemoglobin, *Mtb* resistance pattern, smear status, baseline time to positivity and diabetes mellitus. |
| **Follow-up period** | Starts at randomization (start of treatment) and ends at the occurrence of death or 1 year of follow-up, whichever occurs first. | Starts on the day of starting treatment and ends at the occurrence of death or 1 year of follow-up, whichever occurs first. |
| **Outcome** | Death in the first year after the start of treatment. | Same. |
| **Causal estimand** | Intention-to-treat effect: the average treatment effect of initiating a bedaquiline-based regimen compared to a non-bedaquiline-based regimen. | Observational analogue of the intention-to-treat effect. |
| **Analysis plan** | Intention-to-treat analysis: comparing the probabilities of mortality under each assigned strategy through ratios. | The same intention-to-treat effect with the exception that the re-weighted population is used to estimate the marginal effect using a marginal structural logistic model. |

*Definition of abbreviations:* RR-TB = Rifampicin Resistant Tuberculosis; EXIT-RIF = Evaluating the impact of Xpert on Tuberculosis-RIFampicin Resistance; SMARTT = Sequencing Mycobacteria and Algorithm-determined resistant tuberculosis treatment trial; DAG = Directed Acyclic Graph; HIV = Human Immunodeficiency Virus; *Mtb* = *Mycobacterium tuberculosis*.

For both arms, the follow-up in the emulation trial starts on the day of starting treatment, which is time zero. Random assignment was emulated by inverse probability of treatment weighting (IPTW) using baseline patient characteristics identified by a directed acyclic graph (DAG) of the causal question (17, 18). The outcome of the emulation trial was identical to that of the target trial. To evaluate the causal effect, we estimated the observational analogue of the intention-to-treat effect using the pseudo-population created using IPTW using a marginal structural logistic model.

We followed the Strengthening the Reporting of Observational Studies in Epidemiology (STROBE) guidelines to report this study (19) and The Treatment And Reporting of Missing data in Observational Studies (TARMOS) framework to report missing data (20).

### 2.3. Statistical analysis

To create the pseudo-population in which baseline covariates are independent of the treatment strategy a patient received, we estimated the IPTWs by regressing baseline characteristics against the treatment strategy a patient initiated using logistic regression. Based on a DAG of the causal question (**Figure 1**), we included age, sex, haemoglobin as an indicator of anaemia, weight, diabetes mellitus, history of TB treatment, Human Immunodeficiency Virus (HIV) status, smear status and baseline time to culture positivity as indicators of baseline mycobacterial load, and *Mycobacterium tuberculosis* (*Mtb*) resistance pattern in the model. We used stabilized weights to reduce the variability that can result from extreme weights (17).

**Figure 1:**
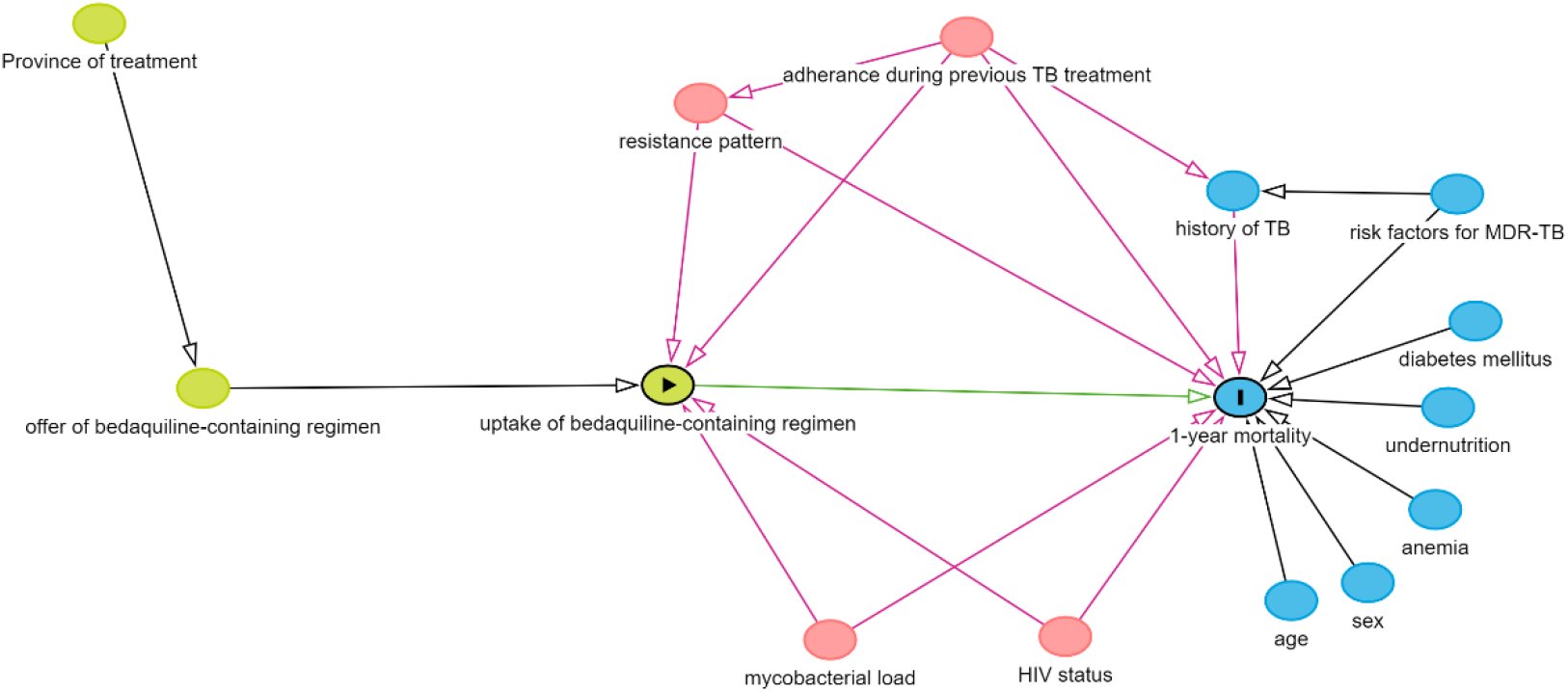
Directed Acyclic Graph used to guide the selection of variables included in the estimation of the inverse probability of treatment weights. *Definition of abbreviations:* HIV = Human Immunodeficiency Virus, TB = Tuberculosis. The green circle with an arrow inside represents the exposure (treatment) in this analysis. The blue circle with a line inside represents the outcome. Red circles are parents of the exposure and outcome and thus confound the exposure outcome-relationship. Blue circles are the parents of the outcome. Green circles are the parents of the exposure.

Age, haemoglobin, weight, and baseline time to culture positivity were modelled as continuous variables in the model, whereas sex, diabetes mellitus, and smear status were included as binary variables. HIV status was modelled as a categorical variable with four levels; HIV negative, controlled HIV (HIV positive, on antiretroviral therapy (ART) and viral load <= 1000 copies/mL or CD4 >= 350 cells/mm^3^), uncontrolled HIV (HIV positive, on ART and viral load >1000 copies/mL or CD4 < 350 cells/mm^3^) and HIV-positive not-on ART. For the *Mtb* resistance pattern, we balanced the groups on resistance to isoniazid and fluoroquinolones as DST on other drugs is not often assessed in the routine management of RR-TB.

Missing data ranged from 3·85% to 24·5%. We theorized the missing data mechanism to be missing at random (MAR), with missingness depending on observed outcomes and/or the covariates (21). We employed multiple imputation using multivariate imputation by chained equations to deal with the missing data. All variables to be included in the IPTW estimation were included in the multiple imputation model, as well as auxiliary variables such as province, prior adherence, baseline culture result, risk factors for RR-TB, HIV status (positive or negative), viral load, CD4 count, and ART status. Multiple imputation was performed separately for patients initiated on a bedaquiline-containing regimen and those initiated on a non-bedaquiline-containing regimen, to ensure that the imputed data accurately reflect the distribution of the true data within each treatment strategy (22).

Based on the maximum number of missingness (24·5%) we did 25 imputations (23). We created the IPTW in the imputed datasets using the ‘within’ method in the MatchThem package in R (24) by creating IPTWs and estimating the treatment effect in each imputed dataset and pooling the treatment effect estimated from each imputed dataset to obtain the overall effect. This method has been shown to be unbiased under the MAR mechanism and accurately considers the variability in each imputed dataset (25). Using the pseudo-populations from the imputed datasets, we then estimated the causal effect using a marginal structural logistic model. We used robust standard errors to calculate the 95% confidence interval for the effect estimate to account for the homogeneity introduced by weighting. All analyses were run in R 4.4.0.

### 2.4. Causal identification conditions

Causal inference from observational data requires investigators to assess that four key assumptions hold: temporality, consistency, exchangeability and positivity. When using IPTW, an additional assumption, no misspecification of the propensity score model (18), should also hold. Temporality requires that the exposure (treatment) must precede the outcome in all participants with events only counted after the exposure (26). Consistency requires that among those treated, their outcome would not have been any different if they had been randomized to that treatment in the target trial.

Similarly, among those untreated, their outcome would not have been any different if they had been assigned to not being treated in the target trial (26). For this assumption to hold, the treatment strategies must be concisely defined and identified in the data. Exchangeability requires that those treated have the same average pre-treatment risk of the outcome as those who were not treated, such that in a randomized trial, assignment to either treatment strategy can be swapped without any impact on the effect estimate (26). Observational studies aim to achieve conditional exchangeability of prognostically important patient characteristics. When using IPTWs, this assumption is valid when the standardized differences between covariates used to estimate the IPTW do not exceed 10%. A standardized difference of more than 10% is indicative of a significant imbalance between treated and untreated groups (27). This assumption can be assessed quantitatively by the standardized differences and visualized on Love plots (28). The assumption of conditional exchangeability can also be assessed by examining the distribution of covariates, which should be equal in those treated and those untreated. Positivity, when using IPTWs, requires that there is a positive (non-zero) probability of receiving each treatment for every stratum defined by exposure and covariates that occur among individuals in the population (26). This can be assessed qualitatively by tabulating the distribution of covariates in each treatment group. This assumption is also tied with the assumption of no misspecification of the propensity score model, whereby if the mean of the stabilized weights deviates from one or the presence of extreme values potentially indicates non-positivity or misspecification of the propensity score model (18).

### 2.5. Sensitivity analysis

Because mortality in RR-TB often occurs during the first few months of treatment (29) we performed a sensitivity analysis to explore whether a bedaquiline-containing regimen has a beneficial effect on early mortality by exploring its effect on mortality at six months.

## 3. Results

### 3.1. Participant selection and overview of the study population

In the EXIT-RIF study, 75 of the 502 patients diagnosed with RR-TB by the Xpert assay were not eligible for inclusion in the emulation trial because they never started RR-TB treatment. Five of the 200 patients in the SMARTT pragmatic trial were not diagnosed by the Xpert assay (**Figure 2**).

**Figure 2:**
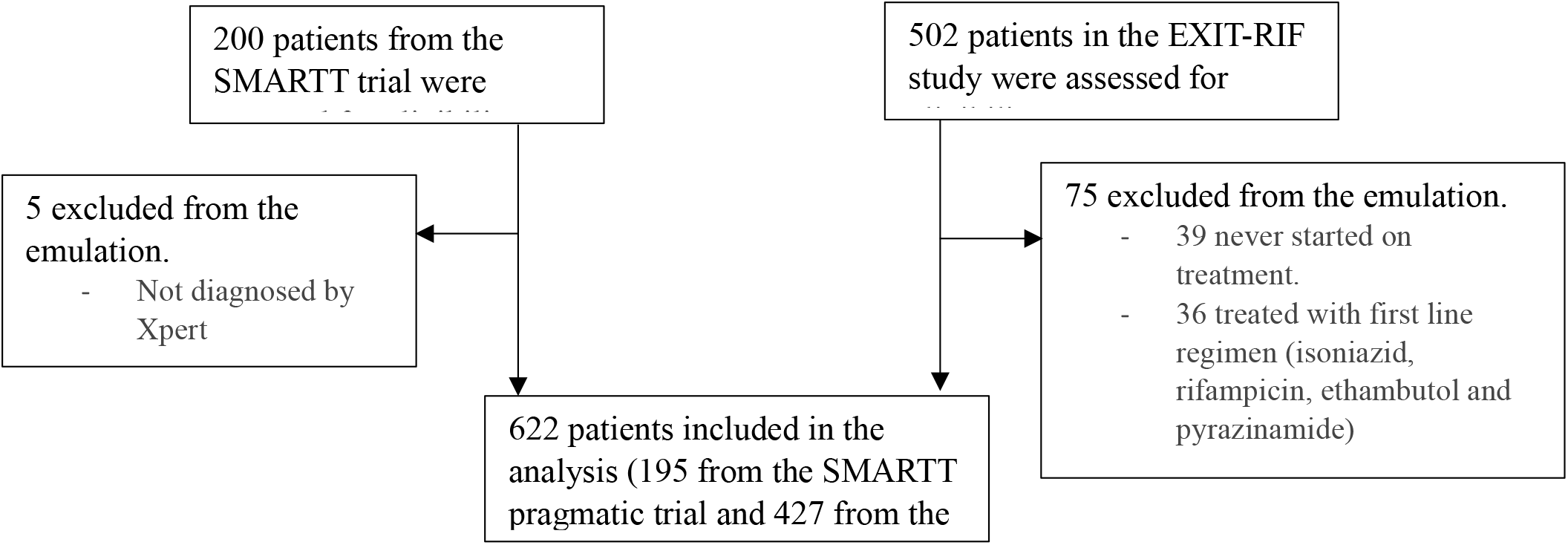
Assessment of eligibility of patients for inclusion in the emulation. Definition of abbreviations: SMARTT = Sequencing Mycobacteria and Algorithm-determined resistant tuberculosis treatment trial, EXIT-RIF = Evaluating the impact of Xpert on Tuberculosis-RIFampicin Resistance

The 622 patients included in the analysis (195 treated with a bedaquiline-containing regimen and 427 treated with a non-bedaquiline-containing regimen) had a mean age of 39 years, were predominantly male, (57%), HIV positive (73%) and 37% had a history of TB treatment (**Table 2**). At diagnosis, most were smear microscopy negative (60%) and the mean time to culture positivity was 12.2 days. Resistance to isoniazid was diagnosed in 36% and resistance to fluoroquinolones in 6.1%. DST for isoniazid was not done in 35% and 59% of the patients did not have results for fluoroquinolone DST. Those initiated on a bedaquiline-containing regimen were more likely to be older, male or have diabetes mellitus.

**Table 2:** Characteristics of the 622 patients included in the emulation of the comparative effectiveness of bedaquiline-containing regimen compared to non-bedaquiline-containing regimen on one-year mortality.

| Variable | Overall, N = 622 | RR-TB Regimen Initiated |  | % Missing |
| --- | --- | --- | --- | --- |
|  |  | BDQ, N = 195 | Non-BDQ, N = 427 |  |
| Age (mean(SD)) | 39.45 (12.23) | 41.98 (12.99) | 38.29 (11.70) |  |
| **Sex** |  |  |  |  |
| Female | 266 (43%) | 58 (30%) | 208 (49%) |  |
| Male | 356 (57%) | 137 (70%) | 219 (51%) |  |
| **Weight (kgs) (mean(SD))** | 53.45 (12.53) | 51.77 (11.59) | 54.38 (12.94) |  |
| (Missing) | 78 |  | 78 | 12.54% |
| **Haemoglobin (g/dL)** | 10.87 (2.60) | 11.36 (2.40) | 10.61 (2.66) |  |
| (Missing) | 104 | 13 | 91 | 16.72% |
| **HIV status** |  |  |  |  |
| HIV negative | 163 (27%) | 71 (40%) | 92 (22%) |  |
| Controlled HIV | 79 (13%) | 49 (28%) | 30 (7.1%) |  |
| Uncontrolled HIV | 114 (19%) | 24 (14%) | 90 (21%) |  |
| HIV positive, not on ART | 242 (40%) | 33 (19%) | 209 (50%) |  |
| (Missing) | 24 | 18 | 6 | 3.85% |
| **Diabetes Mellitus (% Yes)** | 30 (4.8%) | 14 (7.2%) | 16 (3.7%) |  |
| **Smear status** |  |  |  |  |
| Negative | 349 (60%) | 133 (74%) | 216 (54%) |  |
| Positive | 230 (40%) | 47 (26%) | 183 (46%) |  |
| (Missing) | 43 | 15 | 28 | 6.91% |
| **History of TB treatment** |  |  |  |  |
| No history | 359 (59%) | 109 (56%) | 250 (61%) |  |
| Previous First Line | 228 (38%) | 80 (41%) | 148 (36%) |  |
| Previous Second Line | 19 (3.1%) | 5 (2.6%) | 14 (3.4%) |  |
| (Missing) | 16 | 1 | 15 | 2.57% |
| **Isoniazid resistance** |  |  |  |  |
| Sensitive | 184 (30%) | 82 (42%) | 102 (24%) |  |
| Resistant | 223 (36%) | 61 (31%) | 162 (38%) |  |
| Not done | 215 (35%) | 52 (27%) | 163 (38%) |  |
| **Fluoroquinolone resistance** |  |  |  |  |
| Sensitive | 218 (35%) | 105 (54%) | 113 (26%) |  |
| Resistant | 38 (6.1%) | 17 (8.7%) | 21 (4.9%) |  |
| Not done | 366 (59%) | 73 (37%) | 293 (69%) |  |
| **Time to culture positivity (days)** | 12.22 (9.49) | 13.19 (6.91) | 11.79 (10.43) |  |
| (Missing) | 153 | 49 | 104 | 24.59% |
| **Province of treatment** |  |  |  |  |
| Free State | 310 (50%) | 195 (100%) | 115 (25%) |  |
| Gauteng | 129 (21%) |  | 129 (30%) |  |
| Western Cape | 183 (29%) |  | 183 (43%) |  |

*Definition of abbreviations:* RR-TB = Rifampicin-Resistant Tuberculosis, BDQ= Bedaquiline-containing regimen, Non-BDQ = Non-Bedaquiline-containing regimen, SD = standard deviation, kgs = kilograms, g/dL = grams per decilitre, HIV= Human Immunodeficiency Virus, ART = Antiretroviral Therapy, TB = Tuberculosis

### 3.2. Diagnostics of the multiple imputation

Those with complete data differed from those with missing data in terms of smear status, isoniazid resistance and fluoroquinolones resistance (**Supplementary Table 1**). Visualization of the missingness pattern showed groups that differed from each other in missingness, but the missingness was random within the groups defined by missingness (**Supplementary Figure 1**). This observation and an understanding of the causal structure of missingness indicated the plausibility of the MAR assumption. All imputed values were plausible as they were similar to the observed values on examination using strip plots.

### 3.3. Assessment of assumptions for causal inference

By emulating the target trial, we ensured that only eligible patients; those diagnosed by Xpert assay and started on RR-TB treatment, were included in the analysis. Only deaths occurring after the start of treatment (T_0_) were counted as events thus ensuring temporality.

Consistency requires that the causal contrasts of initiation of a bedaquiline-containing regimen compared to initiation of a non-bedaquiline-containing regimen be identified in the data. A review of the data on individual drug administration (start and stop dates) confirmed that all patients from the SMARTT trial initiated a bedaquiline-containing regimen and similarly, all patients from the EXIT-RIF study initiated a non-bedaquiline-containing regimen. Furthermore, all the patients from the SMARTT trial continued to receive bedaquiline after their individualization and none of the EXIT-RIF patients received bedaquiline after their individualization.

To assess conditional exchangeability, we examined the absolute standardized differences for all the variables that were included in the IPTW model before and after weighting, for all the 25 imputations and their distribution (**Figure 3, Supplementary Table 2, Supplementary Figures 3 to 7**). The maximum standardized difference for weight and baseline TTP between the patients initiated on a bedaquiline-containing regimen and those initiated on a non-bedaquiline-containing regimen was 11%, which is not a substantial exceedance of the 10% threshold. The distribution of the categorical variables included in the IPTW model was also similar on examination using bar plots. The assumption of conditional exchangeability for variables included in the IPTW model was therefore considered valid.

**Figure 3:**
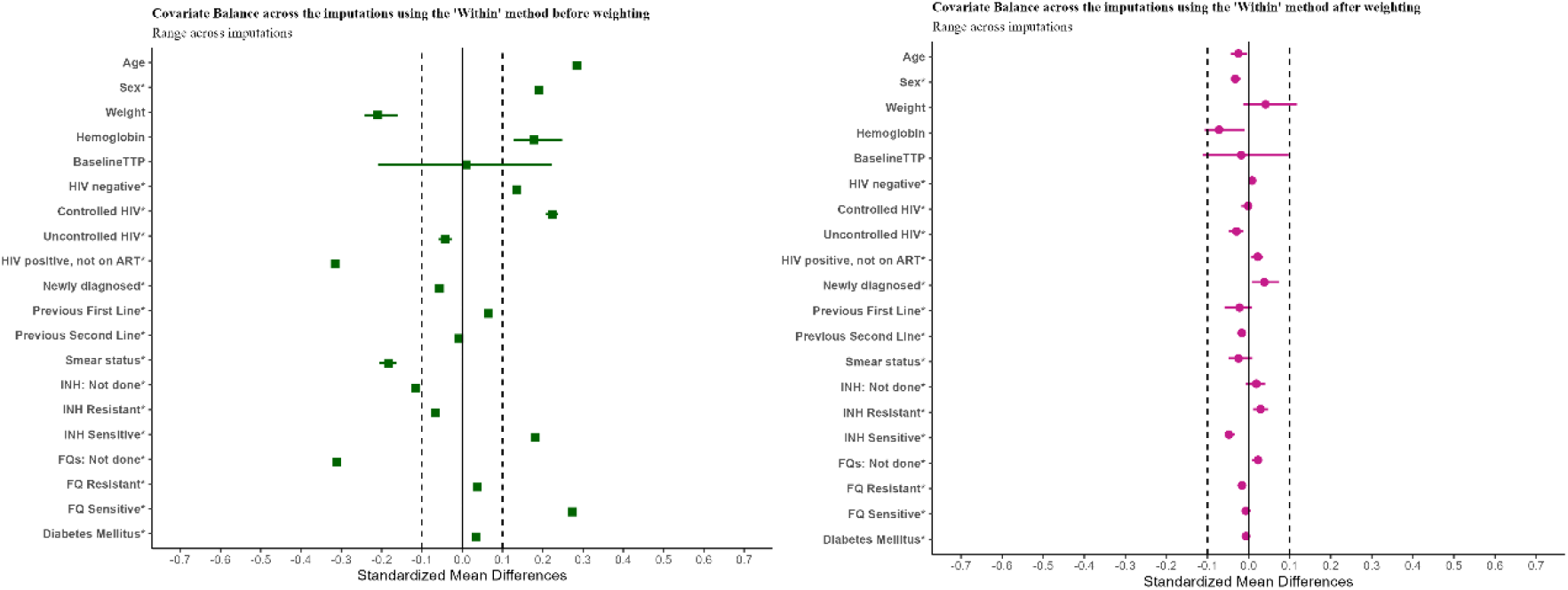
Standardized differences in the covariates. 3A (right): Standardized differences for all the 25 imputations before weighting. 3B: (left) Standardized differences for all the 25 imputations after weighting. *Definition of abbreviations*: TTP = time to culture positivity. HIV = Human Immunodeficiency Virus, ART= Antiretroviral Therapy, INH = Isoniazid, FQ = Fluoroquinolones. *Legend*: 3A shows the standardized differences between the treatment groups before weighting, and 3B shows the standardized differences between the treatment groups after weighting which are all less than 10%, showing that the covariates are balanced in the two treatment groups.

When evaluating the assumptions of positivity and no propensity score model misspecification, we found that the mean of the stabilized weights was 1·007. The distribution of the propensity scores showed good overlap (**Supplementary Figure 8)** and the weights ranged from 0·329 to 14·278 with a standard deviation of 1·03. These results support non-violation of these two assumptions.

### 3.4. Effect estimation

In the original data, one-year mortality was observed in 21% of the patients initiated on a bedaquiline-containing regimen and 34% in the patients initiated on a non-bedaquiline-containing regimen. A logistic regression model estimated 53% lower odds of one-year mortality (OR 0·47, 95% CI: 0·06-0·88) (**Table 3**).

**Table 3:** Effect estimates of initiating a bedaquiline-containing regimen compared to initiating a non-bedaquiline-containing regimen. Effects reported as Odds Ratios and their associated 95% confidence intervals.

| Unadjusted effect<br>(One-year mortality) | Marginal effect<br>(One-year mortality) | Marginal effect on six-month mortality |
| --- | --- | --- |
| 0.47 (0.06 to 0.88) | 0.33 (0.19 to 0.59) | 0.36 (0.20 to 0.63) |

Applying the target trial emulation methodology, a marginal structural logistic model in the re-weighted population, estimated 67% lower odds of one-year mortality had all patients initiated a bedaquiline-containing regimen compared to the counterfactual where all the patients initiated a non-bedaquiline-containing regimen (causal OR 0·33, 95% CI: 0·19-0·59) (**Table 3**).

### 3.5. Sensitivity analysis

The sensitivity analysis found that most of the reduction in mortality caused by a bedaquiline-containing regimen was already achieved in the first six months after initiation of treatment, with a 64% reduction in the odds of mortality among patients who received a bedaquiline-containing regimen compared to a non-bedaquiline-containing regimen (causal OR: 0·36, 95%CI: 0·20-0·63) (**Table 3**).

## 4. Discussion

By emulating a target trial and estimating the average treatment effect, similar to what would be estimated in a randomized clinical trial, our results negate findings of increased mortality in patients treated with bedaquiline reported in phase II (3, 4) and phase III (11) trials and support results of observational studies reporting a reduction in mortality among patients receiving bedaquiline as part of their treatment regimen (12). Our findings are also similar to those reported by an individual patient data meta-analysis that reported a sixty-percent reduction in the odds of mortality among patients treated with bedaquiline, although the data might have included patients from when bedaquiline was offered under compassionate care (30). The effect estimate in our hypothetical trial is larger than that of a similar South African study which found that patients treated with a bedaquiline-containing regimen had an eight per cent lower risk of mortality in the first two years compared to those treated with an injectable-containing regimen (10). We also observed that most of the reduction in mortality occurred during the first six months following initiation of a bedaquiline-containing regimen. This is in contrast to findings from another study in South Africa which did not find a significant effect on six-month mortality (31), which could be because they estimated a conditional effect.

The key strength of our study is that we applied causal inference methods to high-quality observational data to answer a question of public health interest that can no longer be addressed in a randomized clinical trial. Furthermore, we report our methodology and results according to the recent guidance on how to draw causal inferences from observational data (14) (**Supplementary Table 3**). Our methodological approach and the use of the target trial aided the articulation of our question, making the analytical approach transparent and clear. Secondly, the use of IPTW allowed us to use the entire study population in the analysis thus allowing direct interpretability of the findings to the original population. Additionally, we used data where patients were treated under programmatic conditions, adding external validity.

Some limitations of our study include that IPTW relies on the assumption of no unmeasured confounding, which is untestable. We relied on a DAG to determine the prognostically important characteristics that needed to be balanced in the IPTW. However, unaccounted confounding due to misspecification of the DAG cannot be ruled out. The SMARTT trial was performed in the Free State province of South Africa while EXIT-RIF was performed in the Free State, Eastern Cape and Gauteng provinces. Prior studies adjusted for the province of treatment because bedaquiline was rolled out at different times. In the DAG, province is an instrumental variable, affecting the treatment selection process, and thus neither a confounding variable nor a predictor of mortality. Being an instrumental variable, we did not include the province in the IPTW, as this has been shown to increase bias and variance (32). We also encountered some missing data, which was mitigated by multiple imputation.

However, the uncertainty of the missingness could have potentially affected the precision of our estimates. Lastly, our data comes from a high HIV endemic setting thus the findings may not be generalizable to other settings where HIV co-infection is not high.

In conclusion, our results suggest that initiation of a bedaquiline-containing regimen causes a 67% reduction in the odds of one-year mortality. Taken together with the results of other studies, the body of evidence shows that bedaquiline-containing regimens are more effective and reduce mortality compared to other RR-TB regimens that do not contain bedaquiline.

Although countries are increasingly adopting bedaquiline with 92 countries using the shorter 9-month oral regimens and 40 countries reporting to start to use the new 6-month BPaLM/BPaL, by the end of 2022 (1), more access is needed. Future research should investigate whether bedaquiline with the newer backgrounds affords further reduction in mortality.

## Data Availability

All data produced in the present study are available upon reasonable request to the authors.

## References

1. WHO. Global Tuberculosis Report 2023. Geneva: World Health Organization; 2023.

2. FDA. FDA Approval of SIRTURO (bedaquiline): U.S. Food and Drug Administration; December 28, 2012 [Available from: https://www.accessdata.fda.gov/drugsatfda_docs/nda/2012/204384Orig1s000TOC.cfm.

3. Diacon AH, Donald PR, Pym A, Grobusch M, Patientia RF, Mahanyele R, et al. Randomized pilot trial of eight weeks of bedaquiline (TMC207) treatment for multidrug-resistant tuberculosis: long-term outcome, tolerability, and effect on emergence of drug resistance. Antimicrob Agents Chemother. 2012;56(6):3271–6.

4. Diacon AH, Pym A, Grobusch MP, de los Rios JM, Gotuzzo E, Vasilyeva I, et al. Multidrug-resistant tuberculosis and culture conversion with bedaquiline. N Engl J Med. 2014;371(8):723–32.

5. Charan J, Reljic T, Kumar A. Bedaquiline versus placebo for management of multiple drug-resistant tuberculosis: A systematic review. Indian J Pharmacol. 2016;48(2):186–91.

6. Report of the Guideline Development Group Meeting on the Use of Bedaquiline in the Treatment of multidrug resistant tuberculosis, A review of available evidence. 2016 13 March 2017.

7. WHO. WHO consolidated guidelines on drug-resistant tuberculosis treatment. Geneva; 2019.

8. WHO. WHO consolidated guidelines on tuberculosis. Module 4: treatment-drug-resistant tuberculosis treatment, 2022 update. Geneva; 2022.

9. Schnippel K, Ndjeka N, Maartens G, Meintjes G, Master I, Ismail N, et al. Effect of bedaquiline on mortality in South African patients with drug-resistant tuberculosis: a retrospective cohort study. Lancet Respir Med. 2018;6(9):699–706.

10. Ndjeka N, Campbell JR, Meintjes G, Maartens G, Schaaf HS, Hughes J, et al. Treatment outcomes 24 months after initiating short, all-oral bedaquiline-containing or injectable-containing rifampicin-resistant tuberculosis treatment regimens in South Africa: a retrospective cohort study. Lancet Infect Dis. 2022;22(7):1042–51.

11. Goodall RL, Meredith SK, Nunn AJ, Bayissa A, Bhatnagar AK, Bronson G, et al. Evaluation of two short standardised regimens for the treatment of rifampicin-resistant tuberculosis (STREAM stage 2): an open-label, multicentre, randomised, non-inferiority trial. Lancet. 2022;400(10366):1858–68.

12. Tong E, Wu Q, Chen Y, Liu Z, Zhang M, Zhu Y, et al. The Efficacy and Safety of Bedaquiline in the Treatment of Pulmonary Tuberculosis Patients: A Systematic Review and Meta-Analysis. Antibiotics (Basel). 2023;12(9).

13. Hernán MA, Wang W, Leaf DE. Target Trial Emulation: A Framework for Causal Inference From Observational Data. JAMA. 2022;328(24):2446–7.

14. Dahabreh IJ, Bibbins-Domingo K. Causal Inference About the Effects of Interventions From Observational Studies in Medical Journals. JAMA. 2024.

15. De Vos E, Scott L, Voss De Lima Y, Warren RM, Stevens W, Hayes C, et al. Management of rifampicin-resistant TB: programme indicators and care cascade analysis in South Africa. Int J Tuberc Lung Dis. 2021;25(2):134–41.

16. Van Rie A, De Vos E, Costa E, Verboven L, Ndebele F, Heupink TH, et al. Sequencing Mycobacteria and Algorithm-determined Resistant Tuberculosis Treatment (SMARTT): a study protocol for a phase IV pragmatic randomized controlled patient management strategy trial. Trials. 2022;23(1):864.

17. Miguel A. Hernán JMR. Causal Inference: What If. Raton B, editor: Chapman & Hall/CRC; 2020.

18. Cole SR, Hernan MA. Constructing inverse probability weights for marginal structural models. Am J Epidemiol. 2008;168(6):656–64.

19. von Elm E, Altman DG, Egger M, Pocock SJ, Gotzsche PC, Vandenbroucke JP, et al. The Strengthening the Reporting of Observational Studies in Epidemiology (STROBE) statement: guidelines for reporting observational studies. Lancet. 2007;370(9596):1453–7.

20. Lee KJ, Tilling KM, Cornish RP, Little RJA, Bell ML, Goetghebeur E, et al. Framework for the treatment and reporting of missing data in observational studies: The Treatment And Reporting of Missing data in Observational Studies framework. J Clin Epidemiol. 2021;134:79–88.

21. Rubin DB. Inference and missing data. Biometrika. 1976;63(3):581–92.

22. Zhang J, Dashti SG, Carlin JB, Lee KJ, Moreno-Betancur M. Should multiple imputation be stratified by exposure group when estimating causal effects via outcome regression in observational studies? BMC Med Res Methodol. 2023;23(1):42.

23. White IR, Royston P, Wood AM. Multiple imputation using chained equations: Issues and guidance for practice. Stat Med. 2011;30(4):377–99.

24. Farhad Pishgar NG, Clémence Leyrat and Elizabeth Stuart. MatchThem:: Matching and Weighting after Multiple Imputation. The R Journal. 2021;13(2):292–305.

25. Leyrat C, Seaman SR, White IR, Douglas I, Smeeth L, Kim J, et al. Propensity score analysis with partially observed covariates: How should multiple imputation be used? Statistical Methods in Medical Research. 2019;28(1):3–19.

26. Westreich D. Epidemiology by design: a causal approach to the health sciences: Oxford University Press; 2019.

27. Austin PC, Stuart EA. Moving towards best practice when using inverse probability of treatment weighting (IPTW) using the propensity score to estimate causal treatment effects in observational studies. Stat Med. 2015;34(28):3661–79.

28. Greifer N. Covariate balance tables and plots: a guide to the cobalt package. Accessed March. 2020;10:2020.

29. Schnippel K, Firnhaber C, Ndjeka N, Conradie F, Page-Shipp L, Berhanu R, et al. Persistently high early mortality despite rapid diagnostics for drug-resistant tuberculosis cases in South Africa. Int J Tuberc Lung Dis. 2017;21(10):1106–11.

30. Collaborative Group for the Meta-Analysis of Individual Patient Data in MDRTBt, Ahmad N, Ahuja SD, Akkerman OW, Alffenaar JC, Anderson LF, et al. Treatment correlates of successful outcomes in pulmonary multidrug-resistant tuberculosis: an individual patient data meta-analysis. Lancet. 2018;392(10150):821–34.

31. Mohr-Holland E, Daniels J, Reuter A, Rodriguez CA, Mitnick C, Kock Y, et al. Early mortality during rifampicin-resistant TB treatment. Int J Tuberc Lung Dis. 2022;26(2):150–7.

32. Myers JA, Rassen JA, Gagne JJ, Huybrechts KF, Schneeweiss S, Rothman KJ, et al. Effects of adjusting for instrumental variables on bias and precision of effect estimates. Am J Epidemiol. 2011;174(11):1213–22.

