## Supplementary for "Comparative Effectiveness Of Bedaquiline On One-Year Mortality In Rifampicin-Resistant Tuberculosis: A Target Trial Emulation"

### Supplementary Materials

### Extent of missingness

The figure below shows the extent of missingness in each variable and the missingness patterns arising from different combinations of missingness patterns.

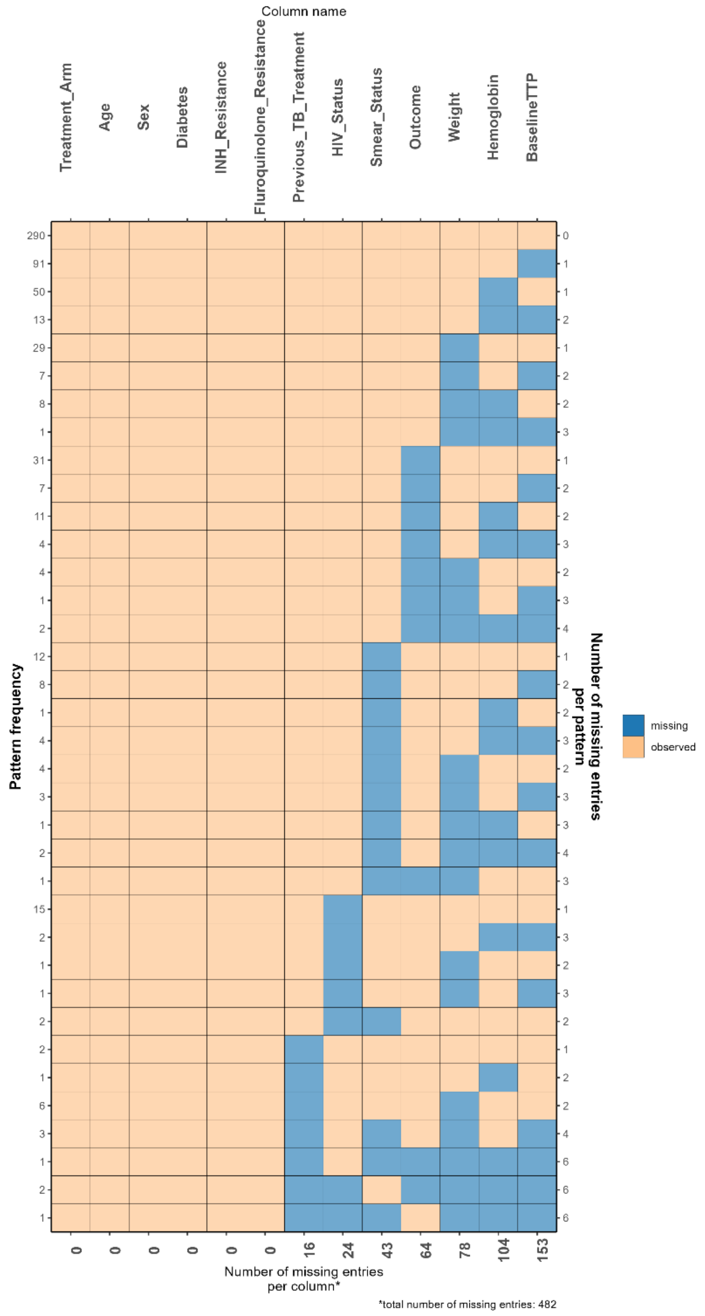


**S Figure 1: Missingness plot showing the number of missing values in each variable and the missingness patterns.** Baseline time to culture positivity had the most missingness because it depends on the culture result, and as such, the culture result was used as an auxiliary variable to aid in its imputation. The most complex missing pattern is in patients who are missing six variables, which was only present in four patients. *Definition of abbreviations*: INH = Isoniazid, TB = Tuberculosis, HIV = Human Immunodeficiency Virus, TTP = Time to culture positivity.

**S Table 1: Characteristics of patients with fully observed data compared to those with missing data.**

| | **Missingness** | | |
|---|---|---|---|
| **Variable** | **Complete**, N = 290[1] | **Missing**, N = 332[1] | **p-value**[2] |
| **Treatment strategy received** | | | 0.2 |
| BDQ | 98 (34%) | 97 (29%) | |
| Non-BDQ | 192 (66%) | 235 (71%) | |
| **Age** (mean(SD)) | 39.03 (12.21) | 39.81 (12.26) | 0.4 |
| **Sex** | | | >0.9 |
| Female | 124 (43%) | 142 (43%) | |
| Male | 166 (57%) | 190 (57%) | |
| **Weight** (mean(SD)) | 53.21 (12.19) | 53.71 (12.92) | 0.9 |
| Unknown | 0 | 78 | |
| **Haemoglobin** | 10.81 (2.51) | 10.95 (2.71) | 0.8 |
| Unknown | 0 | 104 | |
| **HIV status** | | | 0.7 |
| HIV negative | 79 (27%) | 84 (27%) | |
| Controlled HIV | 42 (14%) | 37 (12%) | |
| Uncontrolled HIV | 51 (18%) | 63 (20%) | |
| HIV Positive, not on ART | 118 (41%) | 124 (40%) | |
| Unknown | 0 | 24 | |
| **Diabetes Mellitus (%)** | 12 (4.1%) | 18 (5.4%) | 0.5 |
| **Smear status** | | | 0.045 |
| Negative | 163 (56%) | 186 (64%) | |
| Positive | 127 (44%) | 103 (36%) | |
| Unknown | 0 | 43 | |
| **History of TB** | | | 0.5 |
| No history | 166 (57%) | 193 (61%) | |
| Previous First Line | 113 (39%) | 115 (36%) | |
| Previous Second Line | 11 (3.8%) | 8 (2.5%) | |
| Unknown | 0 | 16 | |
| **INH resistance** | | | <0.001 |
| Not done | 59 (20%) | 156 (47%) | |
| Resistant | 126 (43%) | 97 (29%) | |
| Sensitive | 105 (36%) | 79 (24%) | |
| **FQ resistance** | | | <0.001 |
| Not done | 141 (49%) | 225 (68%) | |
| Resistant | 24 (8.3%) | 14 (4.2%) | |
| Sensitive | 125 (43%) | 93 (28%) | |
| **Time to culture positivity** | 12.26 (8.98) | 12.16 (10.29) | 0.6 |
| Unknown | 0 | 153 | |
| [1]n (%); Mean (SD) | | | |
| [2]Pearson's Chi-squared test; Wilcoxon rank sum test | | | |

*Definition of abbreviations:* SD = Standard deviation, BDQ = Bedaquiline, HIV = Human Immunodeficiency Virus, TB = Tuberculosis, INH = Isoniazid, FQ = Fluoroquinolones

### Sample size

There are no specific guidelines on how sample size or power for observational studies that aim at making causal conclusions should be estimated. Some literature argues that with already available data, power calculations are not necessary (1). To err on the safe side, we calculated a sample size based on a difference in proportion. A recent study on the efficacy of bedaquiline-containing regimens compared to non-bedaquiline-containing regimens in South Africa reported 15.4% and 25.6% mortality in patients treated with bedaquiline-containing regimens and non-bedaquiline-containing regimens respectively (2). With a 90% power and allowing a 5% probability of making type I error, we would require 163 patients treated with a bedaquiline-containing regimen and 163 patients treated with a non-bedaquiline-containing regimen to detect this difference.

### Complete case analysis

In an analysis of the 290 patients who had fully observed information for all the covariates and the outcome, we found that the odds ratio for the causal effect of initiating a bedaquiline-containing regimen compared to a non-bedaquiline-containing regimen was 0.84 (95% CI: 0.77-0.95). Covariate balance and overlap plots for this analysis are shown in **Figures 2 A and B**. This estimate is biased as there were significant differences between the characteristics of patients with fully observed information and those with missing data. This was the motivation for exploring multiple imputation to mitigate this bias.

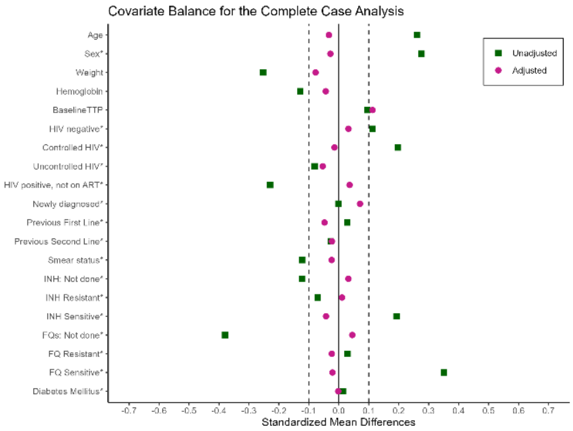


**Figure 2A: Covariate balance for the complete case. This illustrates graphically that conditional exchangeability was achieved.** *Definition of Abbreviations:* Baseline TTP = Baseline Time to culture positivity, HIV = Human Immunodeficiency Virus, ART = Antiretroviral Therapy, INH = Isoniazid, FQ = Fluoroquinolones.

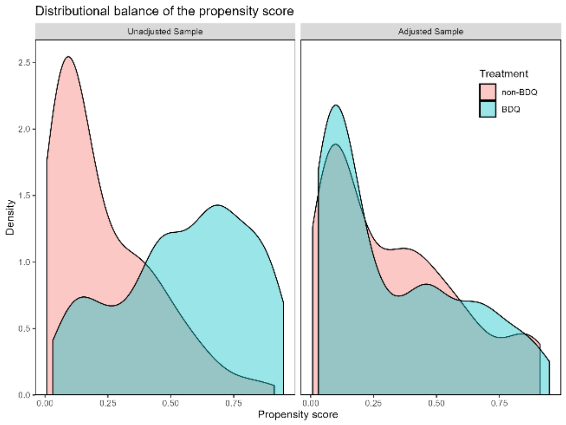


**Figure 2B: Overlap plots for the complete case analysis. There is a good overlap of the propensities for being treated or not treated in both treatment groups** *Legend*: Green hew is for patients treated with bedaquiline-containing regimens and red hew is for patients treated with non-bedaquiline-containing regimen

### Analysis of the multiply imputed data

**Supplementary Table 2** shows the standardized differences of the covariates included in the Inverse probability of treatment weighting (IPTW) model between the patients initiated on a bedaquiline-containing regimen compared to those initiated on a non-bedaquiline-containing regimen.

**S Table 2: Standardized Differences for all the variables used in the Inverse probability of Treatment Weighting (IPTW) model**

| | **Variable modelled as** | **Mean of the Standardized Difference** | **Maximum standardized for all the imputations** | **Mean Kolmogorov Statistic** | **Maximum Kolmogorov Statistic for all the imputations** |
|---|---|---|---|---|---|
| Propensity score | Distance | 0.0254 | 0.0777 | 0.1033 | 0.1305 |
| Age | Continuous | 0.0248 | 0.0439 | 0.0766 | 0.1011 |
| Sex | Binary | 0.0325 | 0.0440 | 0.0325 | 0.0440 |
| Weight | Continuous | 0.0424 | 0.1177 | 0.0747 | 0.0912 |
| Haemoglobin | Continuous | 0.0719 | 0.1073 | 0.1402 | 0.1645 |
| Baseline time to positivity | Continuous | 0.0474 | 0.1111 | 0.3032 | 0.4338 |
| HIV negative | Binary | 0.0085 | 0.0155 | 0.0085 | 0.0155 |
| Controlled HIV | Binary | 0.0045 | 0.0177 | 0.0045 | 0.0177 |
| Uncontrolled HIV | Binary | 0.0295 | 0.0479 | 0.0295 | 0.0479 |
| HIV positive, not on ART | Binary | 0.0222 | 0.0345 | 0.0222 | 0.0345 |
| No history of TB treatment | Binary | 0.0385 | 0.0738 | 0.0385 | 0.0738 |
| Previous First Line | Binary | 0.0227 | 0.0589 | 0.0227 | 0.0589 |
| Previous Second Line | Binary | 0.0165 | 0.0196 | 0.0165 | 0.0196 |
| Smear Positive | Binary | 0.0252 | 0.0495 | 0.0252 | 0.0495 |
| INH Not Done | Binary | 0.0194 | 0.0400 | 0.0194 | 0.0400 |
| INH Resistant | Binary | 0.0292 | 0.0483 | 0.0292 | 0.0483 |
| INH Sensitive | Binary | 0.0480 | 0.0579 | 0.0480 | 0.0579 |
| FQ Not Done | Binary | 0.0230 | 0.0325 | 0.0230 | 0.0325 |
| FQ Resistant | Binary | 0.0161 | 0.0206 | 0.0161 | 0.0206 |
| FQ Sensitive | Binary | 0.0077 | 0.0163 | 0.0077 | 0.0163 |
| Diabetes Mellitus | Binary | 0.0065 | 0.0116 | 0.0065 | 0.0116 |

*Definition of Abbreviations:* HIV = Human Immunodeficiency Virus, ART = Antiretroviral Therapy, TB = Tuberculosis, INH = Isoniazid, FQ = Fluoroquinolones.

### Distributional balance of covariates

**Sex**

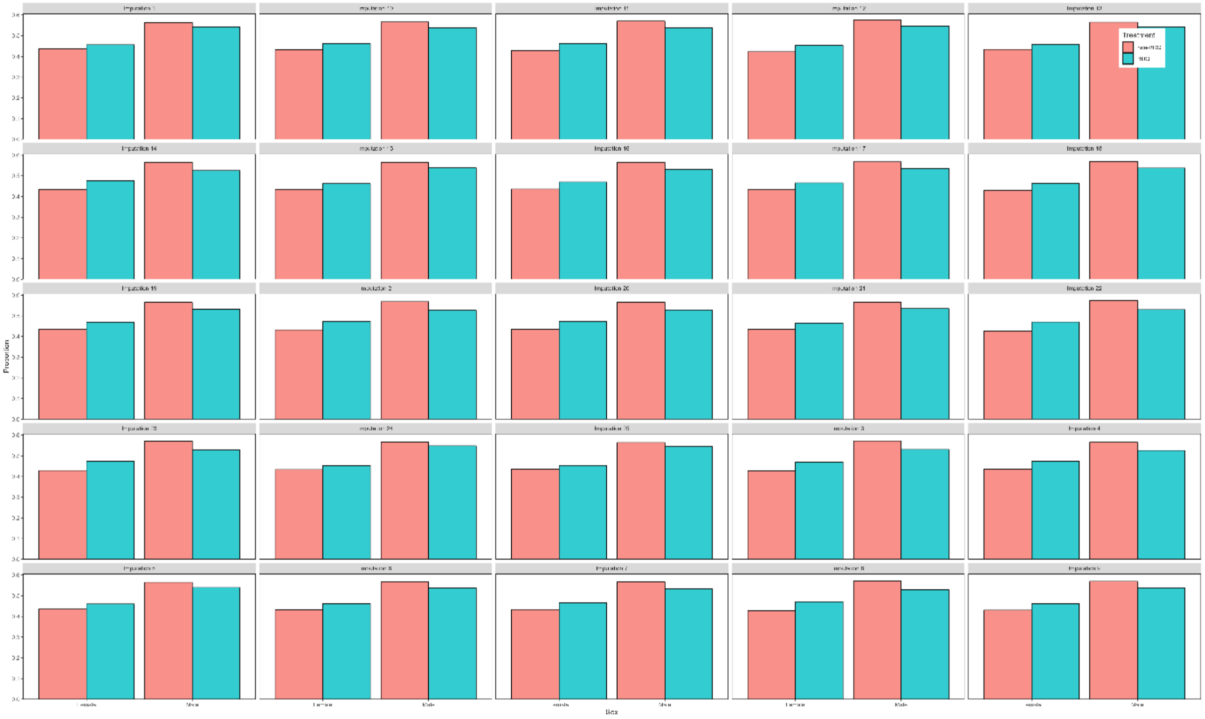

**S Figure 3: Distributional balance for Sex. There are similar proportions of males and females in both treatment groups for all 25 imputations** *Legend*: Green hew is for patients treated with a bedaquiline-containing regimen and red hew is for patients treated with non-bedaquiline-containing regimens.

**Balance for HIV status**

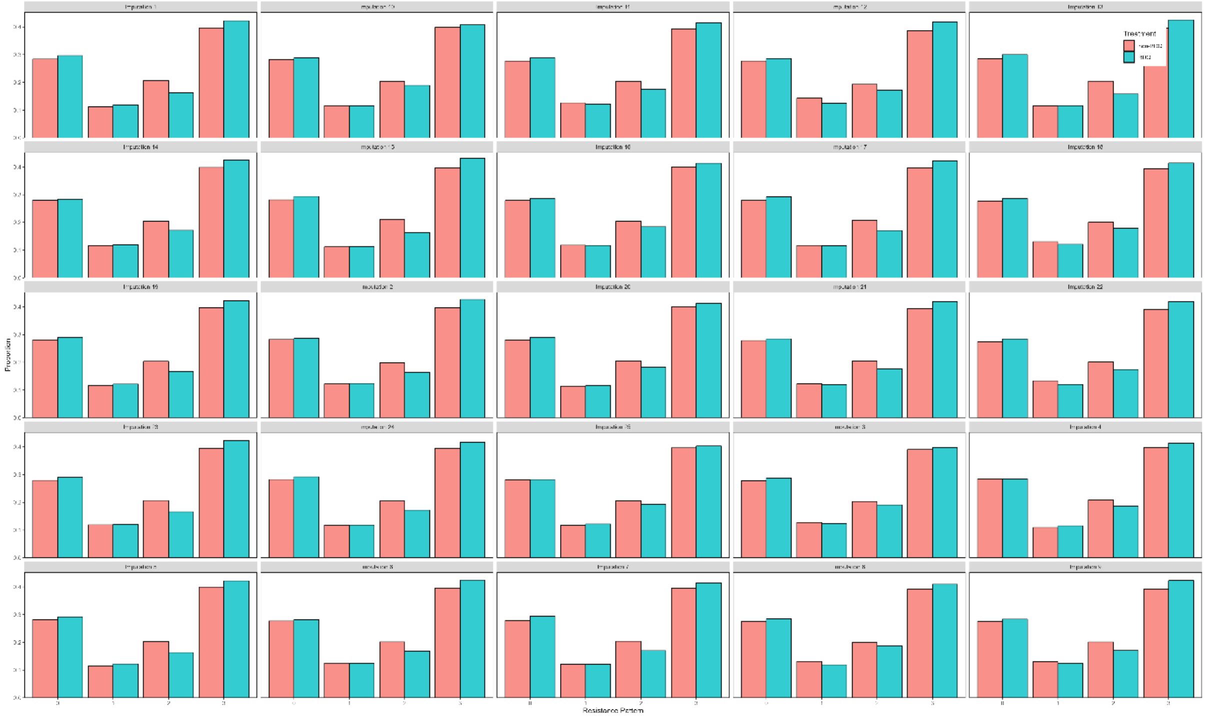


**S Figure 4: Distributional balance for HIV status. There are similar proportions of participants with different HIV statuses in both treatment groups for all the 25 imputations** *Legend*: Green hew is for patients treated with bedaquiline-containing regimens and red hew is for patients treated with non-bedaquiline-containing regimens.

**Balance for History of History of TB Treatment**

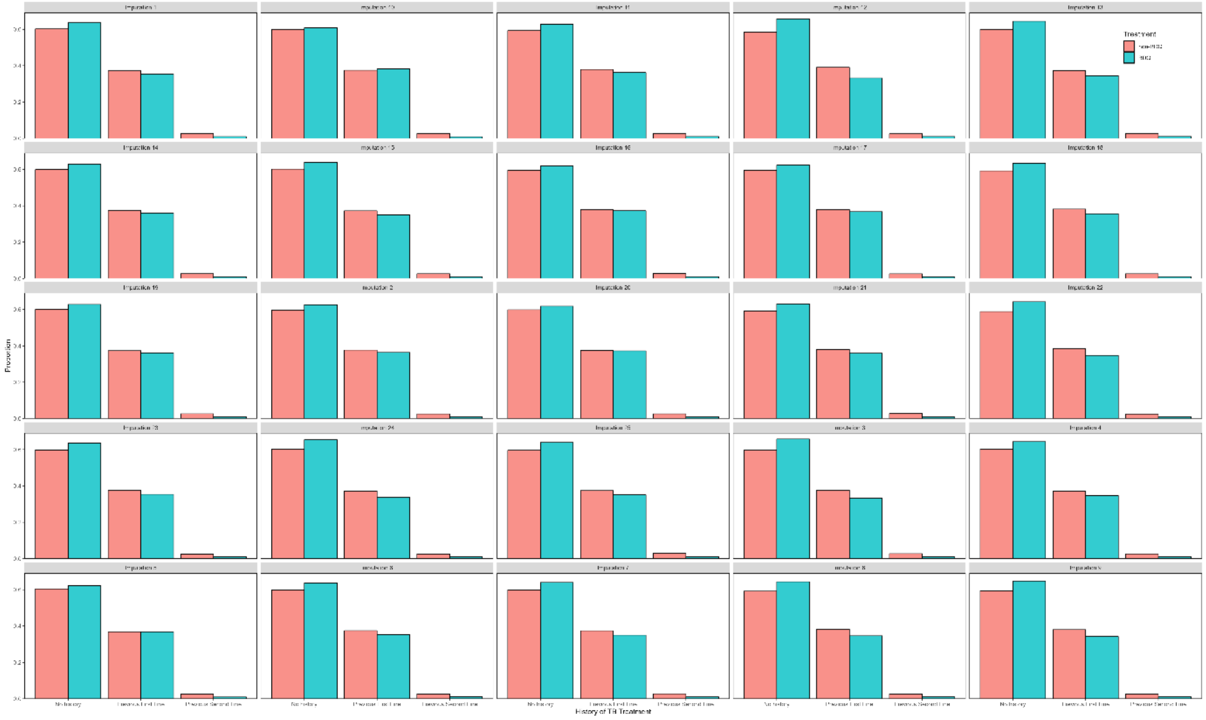

**S Figure 5: Distributional balance for History of TB treatment. There are similar proportions of patients with different histories of treatment in both treatment groups for all the 25 imputations** *Legend*: Green hew is for patients treated with bedaquiline-containing regimens and red hew is for patients treated with non-bedaquiline-containing regimens.

**Balance for INH Resistance**

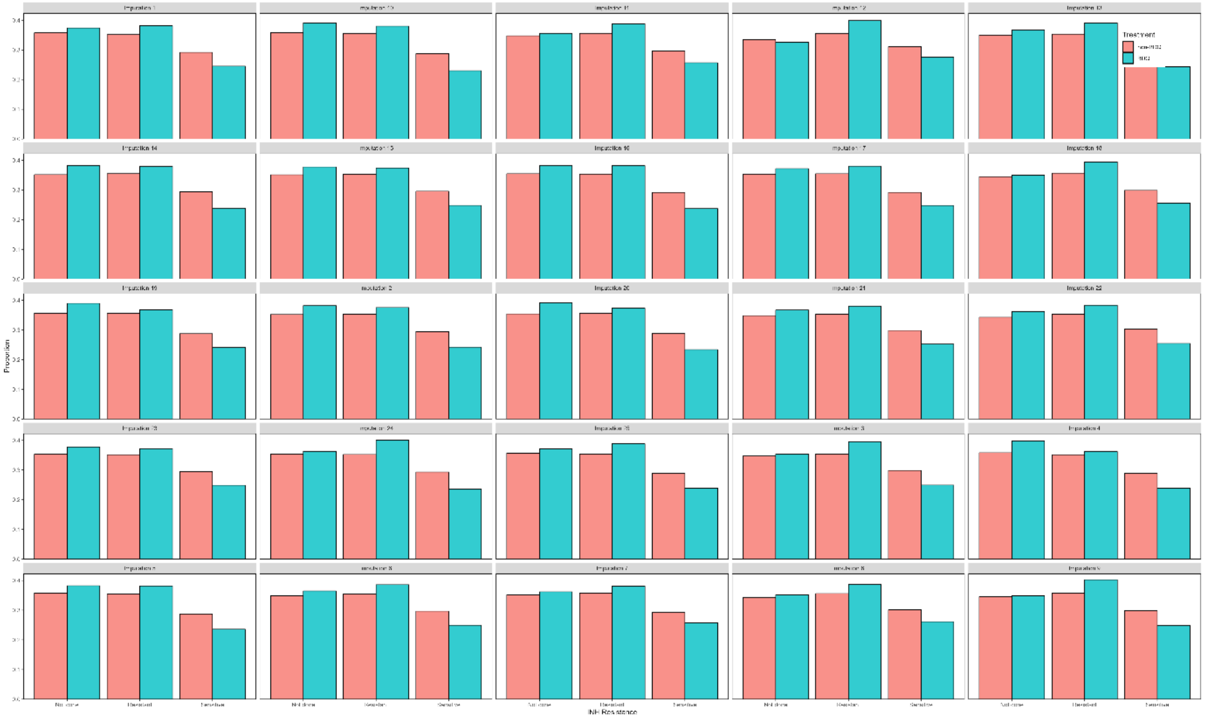

**S Figure 6: Distributional balance for INH resistance. There are similar proportions of patients with different isoniazid statuses in both treatment groups for all the 25 imputations** *Legend*: Green hew is for patients treated with bedaquiline-containing regimens and red hew is for patients treated with non-bedaquiline-containing regimens.

**Balance for FQ Resistance**

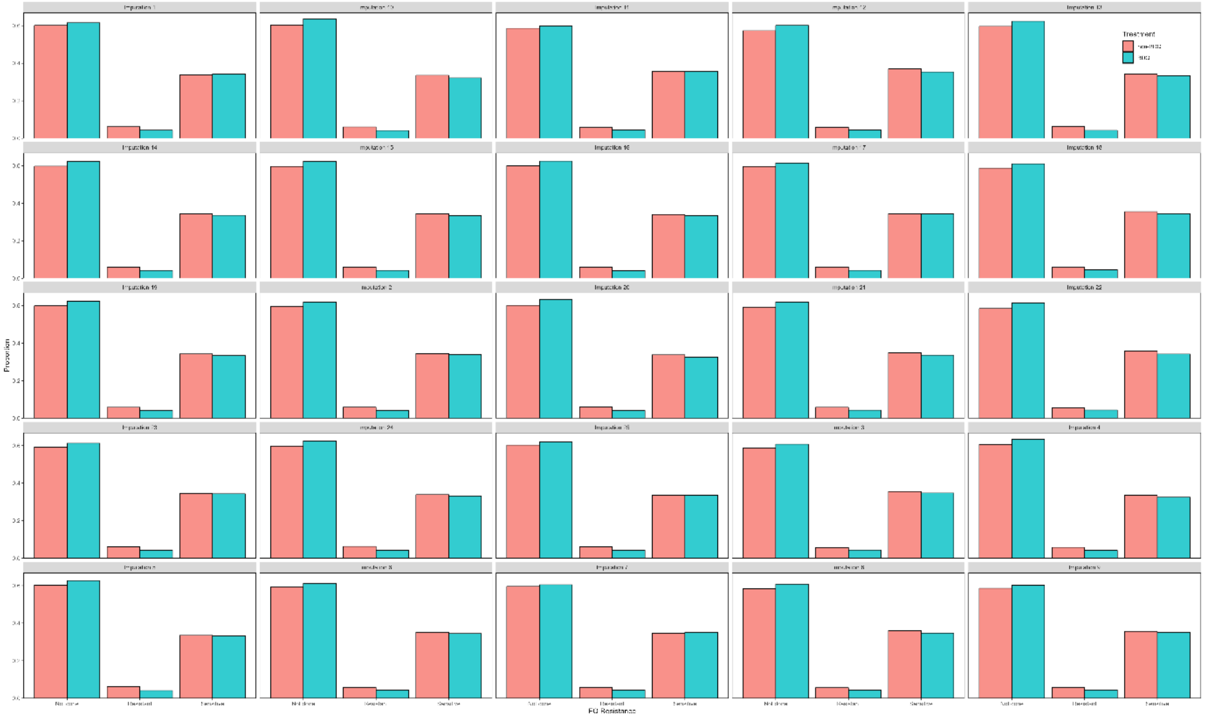

**S Figure 7: Distributional Balance for Fluoroquinolones resistance. There are similar proportions of patients with different fluoroquinolone statuses in both treatment groups for all the 25 imputations** *Legend*: Green hew is for patients treated with bedaquiline-containing regimens and red hew is for patients treated with non-bedaquiline-containing regimens.

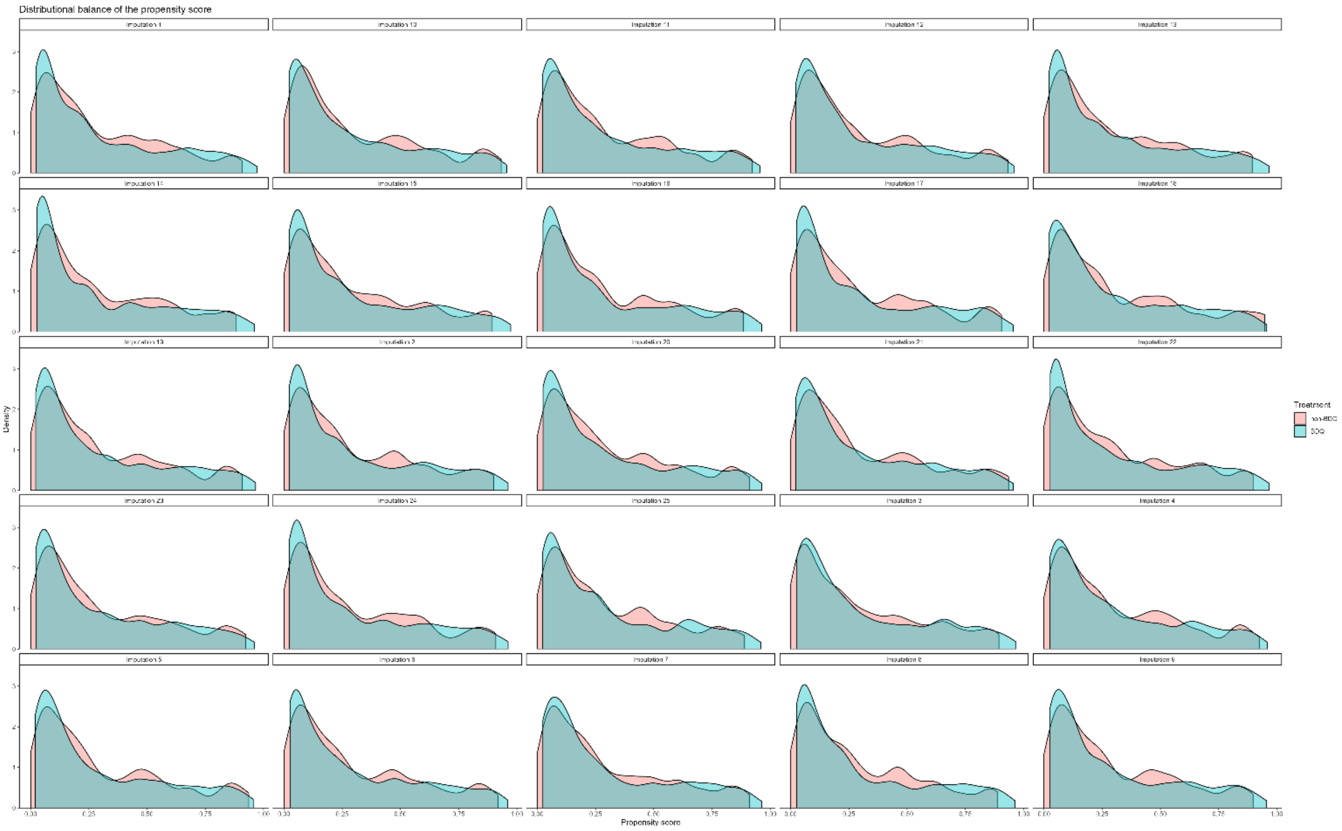


**S Figure 8: Overlap plots for all the imputations. There is a good overlap of the propensities for being treated or not treated in both treatment groups for all the 25 imputations** *Legend*: Green hew is for patients treated with bedaquiline-containing regimens and red hew is for patients treated with non-bedaquiline-containing regimens

**S Table 3: Guiding questions on how to draw causal inferences from observational data. The table shows the questions suggested by (3) and how they were used to guide the analysis.**

| **Question** | **Approach** |
|---|---|
| What is the causal question? | What is the causal effect of initiating a bedaquiline-containing regimen compared to initiating a non-bedaquiline-containing regimen on one-year mortality in RR-TB patients? |
| What quantity would, if known, answer the causal question? | The average treatment effect. |
| What is the study design? | Target trial emulation |
| What causal assumptions are being made? | Temporality, causal consistency, exchangeability, positivity and no misspecification of the propensity score model. |

| | |
|---|---|
| How can the observed data be used to answer the causal question in principle and practice? | 1. Temporality: Events should only be counted after the exposure has been assigned<br>2. Consistency: Among those treated, their outcome would not have been any different if they had been randomized to that treatment in the target trial. Similarly, among those untreated, their outcome would not have been any different if they had been assigned to not being treated in the target trial.<br>3. Exchangeability: Those treated have the same average pre-treatment risk of the outcome as those who were not treated, such that in a randomized trial, assignment to either treatment strategy can be swapped without any impact on the effect estimate.<br>4. Positivity: When using IPTWs, requires that there is a positive (non-zero) probability of receiving each treatment for every stratum defined by exposure and covariate that occur among individuals in the population<br>5. No-misspecification of the propensity score model: The mean of the stabilized weights should not deviate from 1. |
| Is a causal interpretation tenable? | See the results section. |

### References

1. Hernán MA. Causal analyses of existing databases: no power calculations required. J Clin Epidemiol. 2022;144:203-5.
2. Pai H, Ndjeka N, Mbuagbaw L, Kaniga K, Birmingham E, Mao G, et al. Bedaquiline safety, efficacy, utilization and emergence of resistance following treatment of multidrug-resistant tuberculosis patients in South Africa: a retrospective cohort analysis. BMC Infect Dis. 2022;22(1):870.
3. Dahabreh IJ, Bibbins-Domingo K. Causal Inference About the Effects of Interventions From Observational Studies in Medical Journals. JAMA. 2024.
